# Serology characteristics of SARS-CoV-2 infection since the exposure and post symptoms onset

**DOI:** 10.1101/2020.03.23.20041707

**Authors:** Bin Lou, Ting-Dong Li, Shu-Fa Zheng, Ying-Ying Su, Zhi-Yong Li, Wei Liu, Fei Yu, Sheng-Xiang Ge, Qian-Da Zou, Quan Yuan, Sha Lin, Cong-Ming Hong, Xiang-Yang Yao, Xue-Jie Zhang, Ding-Hui Wu, Guo-Liang Zhou, Wang-Heng Hou, Ting-Ting Li, Ya-Li Zhang, Shi-Yin Zhang, Jian Fan, Jun Zhang, Ning-Shao Xia, Yu Chen

## Abstract

**Background:** Timely diagnosis of SARS-CoV-2 infection is the prerequisite for treatment and preventive quarantine. The serology characteristics and complement diagnosis value of antibody test to RNA test needs to be demonstrated.

**Method:** A patient cohort study was conducted at the first affiliated hospital of Zhejiang University, China. Serial plasma of COVID-19 patients and were collected and total antibody (Ab), IgM and IgG antibody against SARS-CoV-2 were detected. The antibody dynamics during the infection were described.

**Results:** The seroconversion rate for Ab, IgM and IgG in COVID-19 patients was 98.8% (79/80), 93.8% (75/80) and 93.8% (75/80), respectively. The first detectible serology marker is total antibody and followed by IgM and IgG, with a median seroconversion time of 15, 18 and 20 day post exposure (d.p.e) or 9, 10 and 12 days post onset, separately. The antibody levels increased rapidly since 6 d.p.o and accompanied with the decline of viral load. For patients in the early stage of illness (0-7d.p.o),Ab showed the highest sensitivity (64.1%) compared to the IgM and IgG (33.3% for both, p<0.001). The sensitivities of Ab, IgM and IgG detection increased to 100%, 96.7% and 93.3% two weeks later, respectively.

**Conclusions:** Typical acute antibody response is induced during the SARS-CoV-2 infection. The serology testing provides important complementation to RNA test for pathogenic specific diagnosis and helpful information to evaluate the adapted immunity status of patient. It should be strongly recommended to apply well-validated antibody tests in the clinical management and public health practice to improve the control of COVID-19 infection.

**Take-Home Message:** Antibody responses are induced after SARS-CoV-2 infection and complement diagnosis value of antibody test to RNA test was observed. Antibody tests are critical tools in clinical management and control of SARS-CoV-2 infection and COVID-19.

## Introduction

In the early December 2019, a novel coronavirus (SARS-CoV-2) was first reported to cause lethality pneumonia in human and person-to-person transmission had been demonstrated soon in Wuhan, the capital city of Hubei province, China.[1] The virus rapidly spread through China and then many other countries globally. Up to March 17, 2020, the virus had resulted in over 190,000 laboratory-confirmed cases of coronavirus disease 2019 (Covid-19) and more than 7800 deaths in 163 countries.[2] The World Health Organization (WHO) has declared Covid-19 a public health emergency of international concern and given a “very high” risk assessment on global level.[3] A recent report from China showed that the median incubation period of Covid-19 infection was 4 days (interquartile range, 2 to 7). [4] Fever, cough and fatigue are the most common symptoms.[1] Severe cases could rapidly progress to acute respiratory distress syndrome (ARDS) and septic shock. The abnormalities on chest computed tomography, particularly the ground-glass opacity and bilateral patchy shadowing, were found in over 80% of patients.[5] Over 80% of patients had lymphopenia, and about 60% of patients had elevated C-reactive protein.[6] However, the clinical and laboratory findings of Covid-19 infection are not distinguishable from pneumonia caused by infection of some common respiratory tract pathogens such as influenza virus, streptococcus pneumoniae and mycoplasma pneumoniae.[7] Hence, the timely diagnosis of SARS-CoV-2 infection is important for providing appropriated medical supports and for preventing spread by quarantining.

Currently, the diagnosis of SARS-CoV-2 infection almost solely depends on the detection of viral RNA using polymerase chain reaction (PCR) based technics.[8] Unfortunately, the sensitivity of RNA test in the real-world is not satisfied, particularly when sample collected from upper respiratory tract is used.[9-12] In Wuhan, the overall positive rate of RNA testing is estimated to be around 30-50% in Covid-19 patients when they come to hospital.[13] Furthermore, the overall throughput of available RNA tests are highly limited by their nature of requiring high workload, needing skillful operators for testing and sample collection, and needing costly instrument and special operate places.[14] As a result, a convenient serological detection is expected to be helpful. However, current knowledge of the antibody response to SAR-CoV-2 infection is very limited. The diagnosis value of antibody test remains to be clearly demonstrated. How many patients would raise antibody response and how long will the antibody convert to be positive since the exposure? Are there any meaningful differences between patients with short and long incubation period? What are the sensitivities of antibody detection for patients in different illness stages? Is there any temporal association between the antibody response and the decline of viral load? In order to answer some of the questions, we investigated the characteristics of antibody responses in 80 Covid-19 patients during their hospitalization periods, through detecting total antibody, IgM and IgG using immunoassays.

## Methods

### Study design and participants

A confirmed COVID-19 case was defined based on the New Coronavirus Pneumonia Prevention and Control Program (6th edition) published by the National Health Commission of China. [15] Briefly, a confirmed case should meet three criteria: 1) fever and/or respiratory symptoms; 2) abnormal lung imaging findings; and 3) positive result of the nucleic acid of SARS-CoV-2. The degree of severity of patient was categorized as critical case if any of the bellowed clinical scenes appeared: 1) with ARDS or oxygen saturation < 93% and needing mechanical ventilation either invasively or non-invasively; 2) shock; and 3) complication of organ functional failure and need intensive care unit support. A Covid-19 patient did not meet the above criteria was defined as non-critical case.

This study enrolls a total of 80 cases of COVID-19, where all patients were admitted to the hospital between Jan 19 and Feb 9, 2020, and were willing to donate their blood samples. All enrolled cases were confirmed to be infected by SARS-CoV-2 through real-time RT-PCR (rRT-PCR) testing. The date of illness onset, clinical classification, RNA testing results during the hospitalization period, and the personal demographic information were obtained from the clinical records. A total of 300 healthy people were enrolled from the local community. This study was reviewed and approved by the Medical Ethical Committee of the first affiliated hospital of Zhejiang University (approval number 2020-IIT-47). Written informed consent was obtained from each enrolled subject.

### Antibody measurement

The total antibody (Ab), IgM antibody and IgG antibody against SARS-CoV-2 in plasma samples were tested using three enzyme linked immunosorbent assays (ELISA-Ab, ELISA-IgM and ELISA-IgG), three colloidal-gold lateral-flow immunoassays (LFIA-Ab, LFIA-IgM and LFIA-IgG) and two chemiluminescence microparticle immunoassays (CMIA-Ab and CMIA-IgM), respectively. The ELISA reagents and LFIA reagents were supplied by Beijing Wantai Biological Pharmacy Enterprise Co., Ltd., China (Beijing, China), and the CMIA reagents were supplied by Xiamen InnoDx Biotech Co., Ltd., China (Xiamen, China). The total antibody detection was based on double-antigens sandwich immunoassay and the IgM antibody detection were based on μ-chain capture immunoassay. Mammalian cell expressed recombinant antigens contained the receptor binding domain (RBD) of the SARS-CoV-2 spike protein were used to develop total antibody and IgM antibody assays. Meanwhile, the IgG antibody kits were indirect immunoassays and a recombinant nucleoprotein of SARS-CoV-2 expressed in *Escherichia coli* was used as coating antigen. All the tests were performed according to the manufacturer’s instructions. Briefly, 100 μL, 50 μL and 20 μL of plasma samples were used for ELISA-Ab, CMIA-Ab and CMIA-IgM respectively, and 10 μL of sample was added to 100 μL of sample diluent buffers for ELISA-IgM. For ELISA-IgG, the sample was 20-fold diluted with 20 mM phosphate buffer (pH 7.0) and 10 μL of dilution buffer was added to 100 μL sample. The measurement processes of ELISA and CMIA were conducted with automatic ELISA analyzer HB-300E (Jiaxing CRED Medical Equipment Co. Ltd., China) and automatic CMIA analyzer Caris 200 (Xiamen UMIC Medical Instrument Co. Ltd., China), respectively. When LFIA-Ab and LFIA-IgM were performed, 10 μL of sample was pipetted onto the sample receiving zone followed by adding two drops of sample buffer. For LFIA-IgG, sample was 1000-fold diluted with sample buffer, then 80 μL of dilution was added onto the sample receiving area. Fifteen minutes later after sample was added, the result of LFIA was observed by eyes and recorded.

### Statistical analysis

Incubation period was defined as interval between earliest date of SARS-Cov-2 exposure and earliest date of symptom onset. Non-normal distribution continue data was described as median with interquartile range (IQR) and compared by Wilcoxon test. Categorical data was summarized as counts and percentages. The 95% confidence intervals of sensitivity and specificity were estimated by binomial exact test. Categorical data was compared using χ2 test or Fisher’s exact test for unpaired proportion, McNemar’s test for paired proportions. Cumulative seroconversion rates were calculated by Kaplan-Meier method. All statistical analysis was conducted by SAS 9.4 (SAS Institute, Cary, NC, USA).

### Patient and public involvement

No patients were involved in setting the research question or the outcome measures, nor were they involved in the design and implementation of the study.

### Dissemination declaration

Dissemination to these groups is not possible/applicable.

## Results

### Characteristics of enrolled COVID-19 patients

Among totally 81 cases of COVID-19 patients admitted in the hospital (before Feb 9, 2020), 80 (99%) patients were enrolled in the study (Table 1). The median age of patients was 55 years (IQR, 45-64 years) and 38.7% were females. The critical patients were significantly older than non-critical patients (p<0.001). The time of SARS-CoV-2 exposure before onset in 45 patients (15 were critical cases) had been determined according to the unambiguous close contact with a confirmed Covid-19 patients through the epidemiological inspection when admission to hospital. The incubation period was ranged from 0 to 23 days with a median of 5 days (IQR, 2 – 10 days). By February 15, a total of 32 patients (40%, all were non-critical cases) were recovered and discharged from hospital and none died.

**Table 1.**
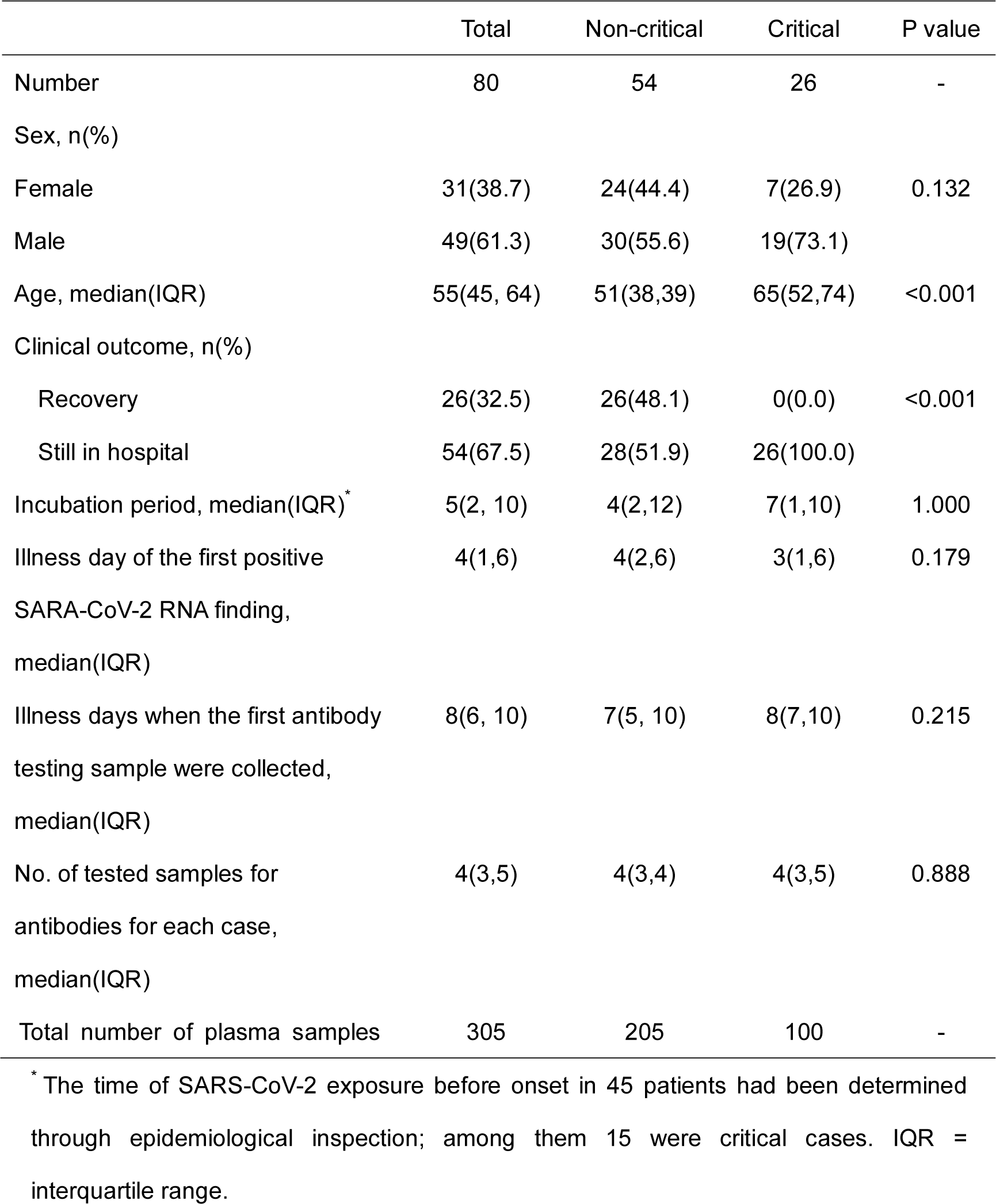
Demographics and clinical characteristics of enrolled COVID-19 patients

### The performance of 2019-nCoV antibody detection kits

A total of 80 Covid-19 patients and 100 to 300 healthy people were tested for antibodies against SARS-CoV-2 using different immunoassays. The seroconversion rate for Ab, IgM and IgG in patients was 98.8% (79/80), 93.8% (75/80) and 93.8% (75/80), respectively (Table 2). The last blood sample of the only patient who had not seroconverted was collected on the 7 days post onset (d.p.o). For Ab, IgM and IgG test, the performance of ELISAs seems the best, although the differences are generally not significant. Therefore, the following serological analyses were all based on the results of ELISAs unless specifically noted.

**Table 2.** Sensitivity and specificity of different immunoassays to detect antibodies against SARS-CoV-2

| Type of immunoassay* | Sensitivity |  |  | Specificity |  |  |
| --- | --- | --- | --- | --- | --- | --- |
|  | No. patient | n(+) | % ( 95%CI ) | No. uninfected person | n(-) | % ( 95%CI ) |
| Total antibody |  |  |  |  |  |  |
| ELISA-Ab | 80 | 78 | 97.5<br>(91.3, 99.7) | 300 | 300 | 100.0<br>(98.8, 100.0) |
| CMIA-Ab | 80 | 77 | 96.3<br>(89.4, 99.2) | 300 | 298 | 99.3<br>(97.6, 99.9) |
| LFIA-Ab | 80 | 78 | 97.5<br>(91.3, 99.7) | 209 | 199 | 95.2<br>(91.4, 97.7) |
| Combined | 80 | 79 | 98.8<br>(93.2, 100.0) | 209 | 197 | 94.3<br>(90.2, 97.0) |
| IgM |  |  |  |  |  |  |
| ELISA-IgM | 80 | 74 | 92.5<br>(84.4, 97.2) | 300 | 300 | 100<br>(98.8, 100) |
| CMIA-IgM | 80 | 69 | 86.3<br>(76.7, 92.9) | 300 | 298 | 99.3<br>(97.6, 99.9) |
| LFIA-IgM | 80 | 71 | 88.8<br>(79.7, 94.7) | 209 | 205 | 98.1<br>(95.2, 99.5) |
| Combined | 80 | 75 | 93.8<br>(86.0, 97.9) | 209 | 203 | 97.1<br>(93.9, 98.9) |
| IgG |  |  |  |  |  |  |
| ELISA-IgG | 80 | 71 | 88.8<br>(79.7, 94.7) | 100 | 100 | 100.0<br>(96.4, 100.0) |
| LFIA-IgG | 80 | 69 | 86.3<br>(76.7, 92.9) | 209 | 208 | 99.5<br>(97.4, 100.0) |
| Combined | 80 | 75 | 93.8<br>(86.0, 97.9) | 100 | 99 | 99.0<br>(94.6, 100) |

\* ELISA, enzyme linked immunosorbent assay; CMIA, chemiluminescence immunoassay;
LFIA, lateral flow immunoassay. The combined sensitivities were calculated based on positive findings by any of the assays, and the combined specificities were calculated based on negative findings for all assays.

The seroconversion was sequential appeared for Ab, IgM and then IgG, with a median onset time of 9, 10 and 12 days, separately (Figure 1A). The seroconversion of Ab was significantly quicker than that of IgM and IgG (p < 0.001). The cumulative seroconversion curve showed that the rate for Ab, IgM reached 100% and IgG reached 97.1% on day 16, 21 and 29 post symptoms onset, correspondingly. The antibody levels increased rapidly since 6 day post onset (d.p.o) (Figure 1B). The decline of viral load co-occurred with the rising of antibody levels. For patients in the early (0-7d.p.o) stage of illness, the ELISA-Ab showed the highest sensitivity (64.1%) compared to the ELISA-IgM and ELISA-IgG (33.3% for both, p<0.001, Table 3). The sensitivities of antibody, IgM and IgG detection increased significantly when the patient entered the later stage and reached 100%, 96.7% and 93.3% two weeks later (p<0.05).

**Table 3.** Performance of different detections in different periods post onset

| Days after onset | No. | RNA* |  | ELISA-Ab |  | ELISA-IgM |  | ELISA-IgG |  |
| --- | --- | --- | --- | --- | --- | --- | --- | --- | --- |
|  | Patients | n(+) | Sensitivity (%) | n(+) | Sensitivity (%) | n(+) | Sensitivity (%) | n(+) | Sensitivity(%) |
| 0-7 | 39 | 36<sup>\$</sup> | 100.0 | 25 | 64.1 | 13 | 33.3 | 13 | 33.3 |
| 8-14 | 75 | 64<sup>\$</sup> | 90.1 | 74 | 98.7 | 65 | 86.7 | 57 | 76.0 |
| 15-29 | 60 | 41<sup>\$</sup> | 70.7 | 60 | 100.0 | 58 | 96.7 | 56 | 93.3 |

\* RNA was tested using deep sputum sample.
\$ There were 36, 71 and 58 patients had RNA testing during the periods between 0-7, 8-14 and 15-29 days post onset, respectively.

**Figure 1.**
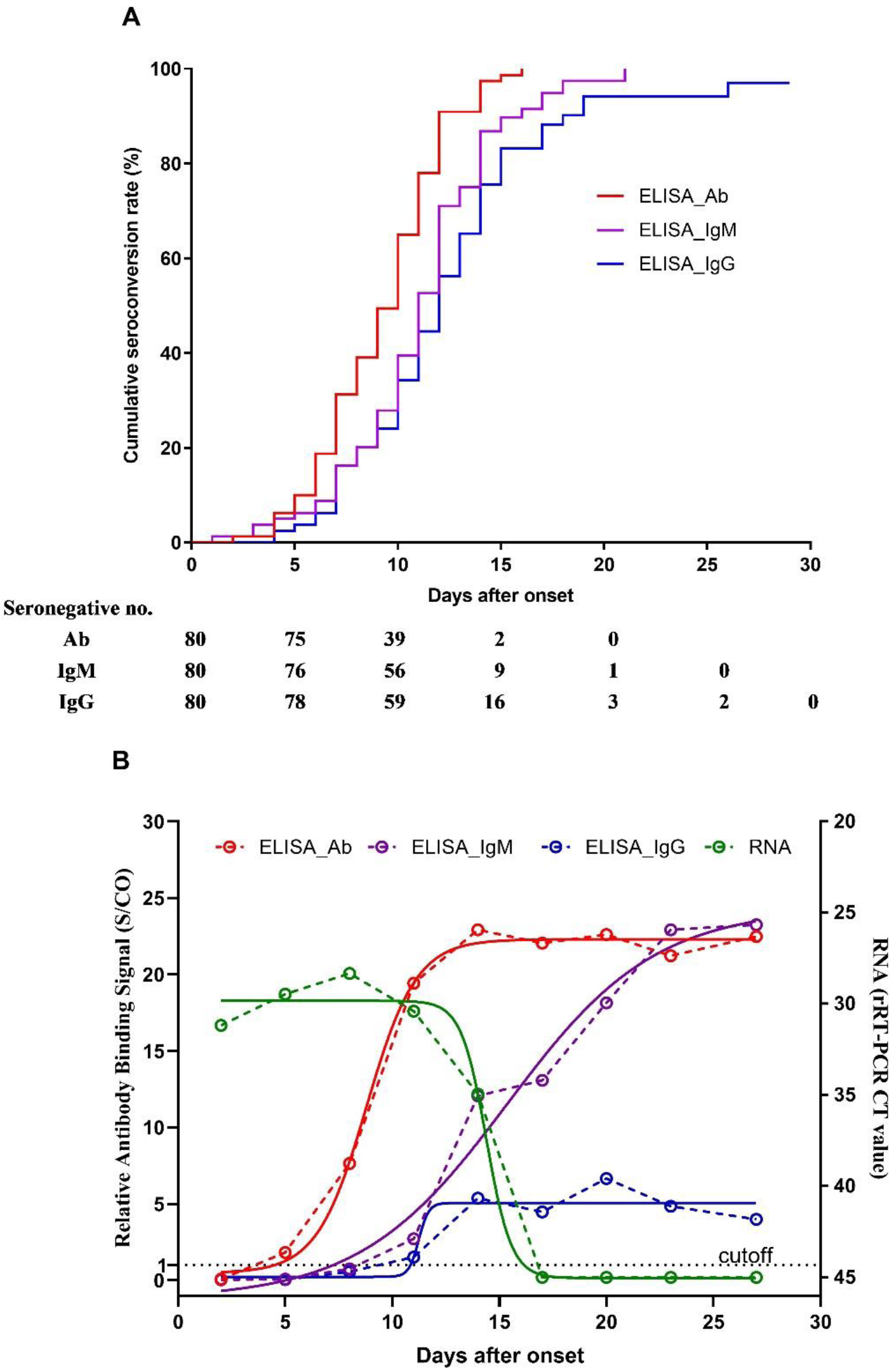
Cumulative seroconversion rates and the dynamics of antibody levels since the onset of illness in 80 Covid-19 patients. (A) The curves of the cumulative seroconversion rates for total antibody, IgM and IgG detected by ELISAs were plotted according to Kaplan–Meier methods. (B) The antibody levels were surrogated expressed using the relative binding signals compared to the cutoff value of each assay (S/CO). Four parameter logistic fitting curves were used to mimic the trends of antibody levels.

### The antibody dynamics after exposure to 2019-nCoV

The antibody dynamics following initial infection were described among 45 patients whose exposure time were determined (Figure 2). The seroconversion was sequential appeared for Ab, IgM and then IgG and the levels increased rapidly, with a median day post exposure (d.p.e) of 15, 18 and 20, separately (Figure 3). The decline of viral load co-occurred with the rising of antibody levels. The cumulative positive rate for Ab, IgM and IgG separately reached 100%, 94.2% and 96.7% at 37 d.p.e. The patients who reported symptoms within 5 days (0-5d) since the exposure were assigned into the short incubation period group and the remaining patients were assigned into the long incubation period group (Table 4). There was no significant difference on age, viral load in the early stage of illness and the risk of critical status between the groups. However, the median seroconversion time was shorter for the short incubation period group than for the long incubation period group (13d.p.e v.s. 21d.p.e, p<0.001). In contrast, the median seroconversion day post onset was longer for the short incubation period group than the long incubation period group (10d.p.o v.s. 7d.p.o, p<0.05).

**Table 4.** Comparison of patients with short or long incubation period

|  | Short incubation<br>period group<br>(0-5d) | Long incubation<br>period group<br>(6-23d) | P value |
| --- | --- | --- | --- |
| Number of patient | 25 | 20 | - |
| Incubation period, median(IQR) | 2(0, 4) | 13(9, 17) | <0.001 |
| Age | 51(36, 63) | 54(46,65) | 0.784 |
| Proportion of critical case (n, %) | 7(28.0) | 8(40.0) | 0.396 |
| Viral load in the early stage of<br>illness, median (IQR)* | 27.8(22.6,34.9) | 27.0(24.1, 30.1) | 0.599 |
| Day post exposure when the<br>antibody converted, median (IQR) | 13(9, 15) | 21(17, 25) | <0.001 |
| Day post onset when the antibody<br>converted, median (IQR) | 10(9, 12) | 7(6, 10) | 0.013 |

\*The viral load was measured using the CT value of real time RT-PCR when detect the first available sputum samples collected after the onset.

**Figure 2.**
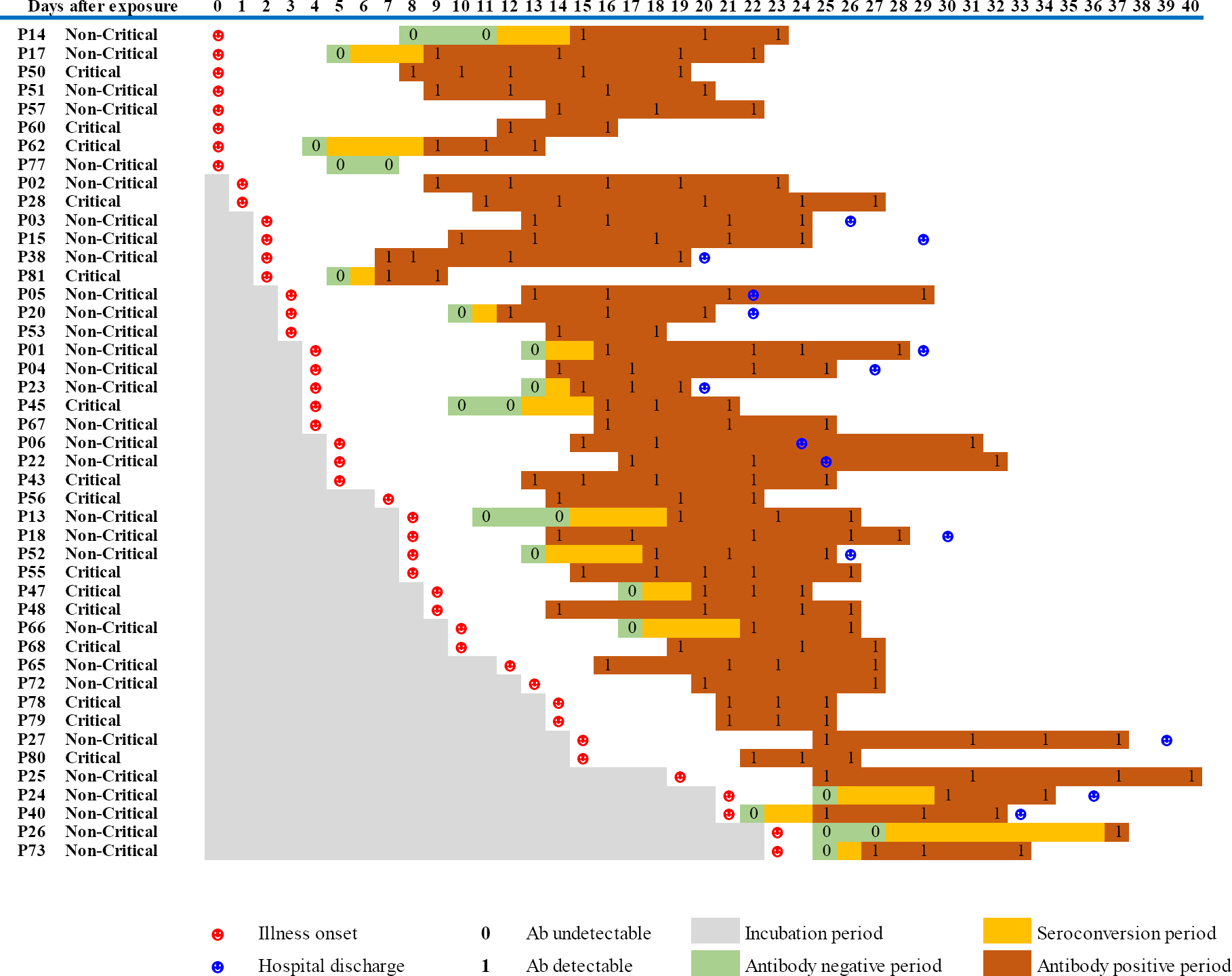
The total antibody dynamics of 45 patients with determined exposure time.

**Figure 3.**
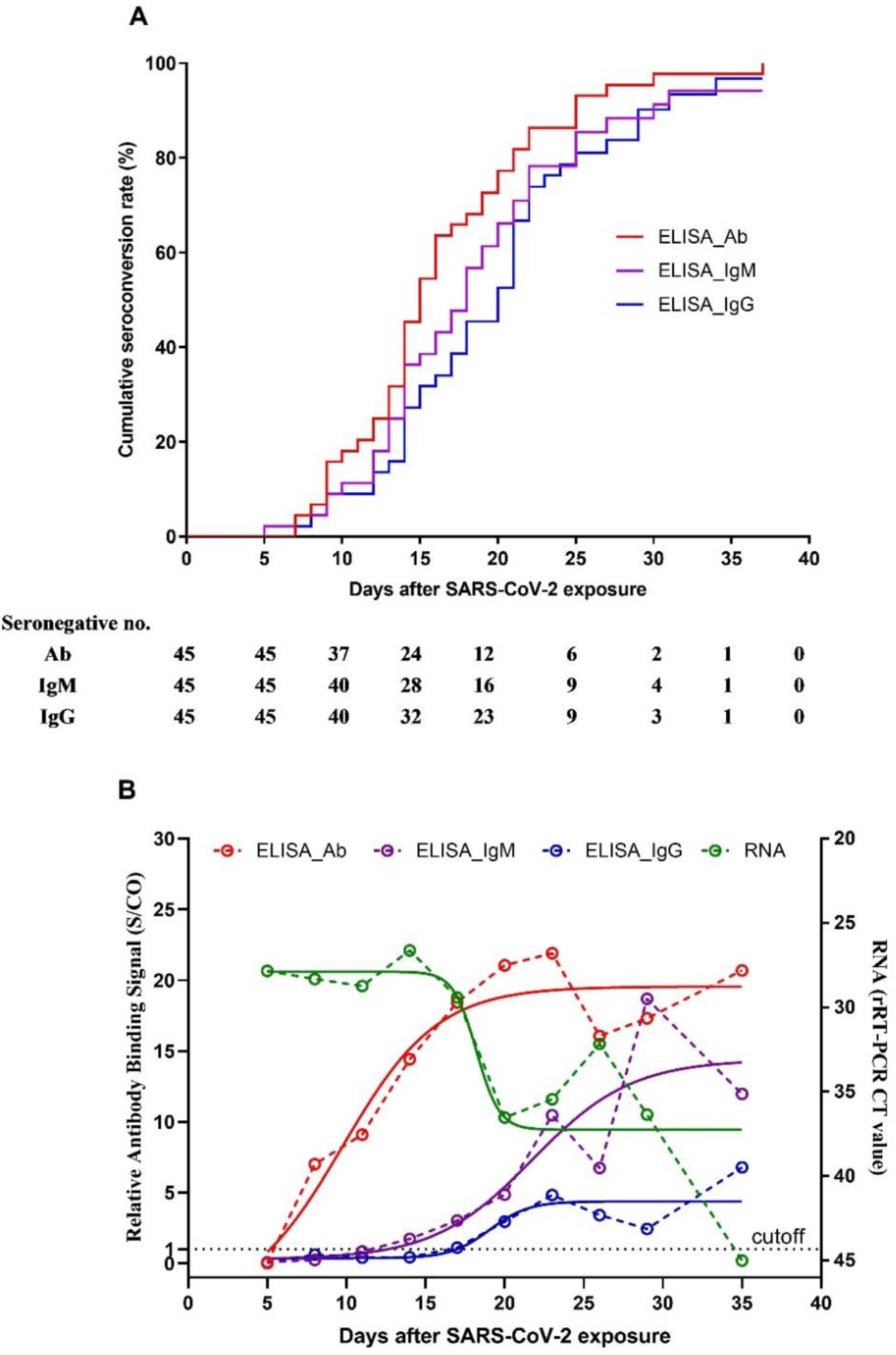
Cumulative seroconversion rates and the dynamics of antibody levels since the exposure of SARS-CoV-2 in 45 Covid-19 patients. (A) The curves of the cumulative seroconversion rates for total antibody, IgM and IgG detected by ELISAs were plotted according to Kaplan–Meier methods. (B) The antibody levels were surrogated expressed using the relative binding signals compared to the cutoff value of each assay (S/CO). Four parameter logistic fitting curves were used to mimic the trends of antibody levels.

## Discussion

The present study showed that near all (98.8%, 79/80) Covid-19 patients converted to be seropositive during the illness course. Seroconversion was first observed on 7d.p.e (2d.p.o). The first detectible antibody is total antibody and followed by IgM and IgG, with a median seroconversion time of 15d.p.e (9d.p.o), 18d.p.e (10d.p.o) and 20d.p.e (12d.p.o). It is very similar as that observed in SARS-CoV-1 infection.[16, 17] Interestingly, the rising of antibody levels accompanied with the decline of viral load. Currently, the quarantine period is set as 14 days since close contacting with a confirmed SARS-CoV-2 infection case. The proportion of infection could not be screened out through RNA test during the quarantine period remains unknown. However, it had been documented that some close contactors presented symptoms and causing transmission after de-isolation[18]. It is believed that antibody test might improve the sensitivity to find infections, but the evidence had not been shown before.

Our data showed that the cumulative seroconversion rate is 45.5% on 14d.p.e. It indicated that about half of the carriers could be screened out if antibody were tested before de-isolation. The cumulative seroconversion rate reached 75% on 20d.p.e (11d.p.o). Seven patients (15.6%) had not present any symptoms before 14 d.p.e, among them 4 patients remained seronegative on 22 d.p.e (1 patient) and 25 d.p.e (3 patients). Hence, follow-up antibody testing and monitoring respiratory symptoms for two weeks after de-isolation would be helpful to further reduce the risk of spread.

The median incubation period was 5 days (IQR, 2 to 10), very similar as previously reported. [4] The needing time for seroconverting since exposure is significantly longer for patients with long incubation period than for patients with short incubation period (21d v.s. 13d, p<0.001). In contrast, the antibody of patients with longer incubation period appeared on the earlier days post onset (7d.p.o v.s. 10d.p.o, p<0.05). The difference seems not be biased by the competence of host immunity because of the similar age distribution between groups. Hence, it is more likely due to the lower intake dose of virus or less efficiency of virus reproduction in the host. Nevertheless, the longer incubation period does not change the initial viral load when the symptoms onset and also the risk of experiencing critical status. Therefore, in addition to the onset time, the interpretation of the negative findings of antibodies in suspected infections should consider the exposure time as well. The faster seroconversion post onset suggests the longer incubation period.

Recently, Guan et al [4] reported that the median time from onset of symptoms to need mechanical ventilator support is 9.3 days. Therefore, timely diagnosis and admit to hospital within 7d.p.o might be crucial to lower the fatality of Covid-19 infection. Zhao et al [19] reported that the overall RNA positive rate was lower than 70% in patients on the first week post symptoms onset and fall to 50% on the next week. Many reports indicated that lots of patients were finally diagnosed by RNA testing through day and day’s repeat sampling and testing, and many cases that were strongly epidemiologically linked to SARS-CoV-2 exposure and with typical lung radiological findings remained RNA negative. [9] In our study, the RNA positive rate is 100% when admitted in hospital, but it cannot be excluded that some patients missed diagnosis due to their undetectable viral RNA. Another reason for the relatively high RNA positive rate in our study is that we used the deep sputum sample for RNA test, in contrast to the more convenient and popular throat/nasal swabs in many other hospitals. It suggests that the lower respiratory samples such as deep sputum and bronchoalveolar lavage might be more reliable for SARS-CoV-2 RNA detection.

Similar as the RNA test, the negative antibody finding in the early stage of illness could not exclude the possibility of infection. Our study showed that the seroconversion rate on 7d.p.o reached 64.1% and then increased to over 90% in the next week. The relative long window period of seroconversion indicates that, for searching previously exposed patients or subclinical carriers, the specific serology should not replace RNA detection but could be an important complement. Furthermore, the fact that near all patients will convert to antibody positive and the titer increased rapidly underlines the usage of antibody test in confirming or excluding diagnosis of SARS-CoV-2 infection if the convalescent sample were tested.

The present data showed that the sensitivity of total antibody detection was higher than that of IgM and IgG (p<0.001) while the specificities are overall comparable when the same testing technic (ELISA, CLMA or LFIA) is used. The detection of total antibody is based on double-antigen sandwich methodology. It can detect any types of antibodies including IgM, IgG and IgA in principle. Besides, it needs the two Fab arms of the same antibody molecular binding to the coated antigen and the enzyme conjugated antigen, which guarantees the specificity of the test and then allows high concentration of antigens be used for coating and second binding to increase the sensitivity of the assay. Therefore, it is not unexpected that the sensitivity of total antibody detection superiors to that of IgM and IgG detection in our study. Usually, IgM is considered as a marker of current or recent infection, while IgG be a marker of post or recent infection. The implications of total antibody are not so straight and then less being used in clinical practice. However, total antibody test outweighs IgM and IgG if the sensitivity is of the top priority, and has been frequently used in blood transfusion products screening such as human immunodeficiency virus and hepatitis C virus. As for current urgently needs of sensitive diagnosis for SARS-CoV-2 infection to contain the spread, Ab test might be a better choice than IgM or IgG, particularly considered the fact that the virus had invaded human society for less than 4 months then the prevalence of antibody induced by post infection is nearly zero.

In our study, antibody tests based on ELISA, CMIA and LFIA were validated and showed good performance overall. CMIA takes the advantages of automatic-operation, rapid and high-throughput, objective and quantitative, but requires costly specific instrument. ELISA is low-cost, objective and high-throughput, wildly used in most of medical laboratories worldwide. LFIA is a rapid point-of-care test, not require special instrument, very convenience and easy to operate but the reading of result is subjective. The establishment of different immunoassays provides flexible choices for the users.

The limitations of the study included: 1) only symptomatic infections were enrolled, therefore if the antibody response to asymptomatic infection follows similar features remains to be determined; 2) most blood samples were all collected with one month post onset so the lasting time of antibodies cannot be estimated; 3) the antibody levels had not been exactly titrated. Future studies are needed to better understand the antibody response profile of SARS-CoV-2 infection and to precisely interpret the clinical meaning of serology findings.

In conclusion, typical acute antibody response is induced during the SARS-CoV-2 infection. The serology testing provides important complementation to RNA test for pathogenic specific diagnosis and helpful information to evaluate the adapted immunity status of patient. It should be strongly recommended to apply well-validated antibody tests in the clinical management and public health practice to improve the control of Covid-19 infection.

## Data Availability

We will share individual participant data that underlie the results reported in this article after deidentification (text, tables, figures and appendices). The data will be available beginning 6 months after the major findings from the final analysis of the study were published, ending 2 years later. The data will be shared with investigators whose proposed use of the data has been approved by an independent review committee identified for individual participant data meta-analysis. Proposals should be directed to. To gain access, data requestors will need to sign a data access agreement.

## Acknowledgements

We acknowledge the work and contribution of all the health providers from the First Affiliated Hospital of Zhejiang University. We sincerely thanked Shan Qiao, Xue-Rong Jia, Bao-Liang Jia, Wen-Jie Sun Ji-Pei Zhang, Shuang Qiu from Beijing Wantai Biological Pharmacy Enterprise Co. Ltd., Liu-Wei Song, Shun-Hua Wen, Rong-Hua Sun, Ji-Pei Shen from Xiamen Innodx Biotech Co. Ltd., Fei-Hai Xu from Xiamen UMIC Medical Instrument Co. Ltd. for their helpful technical assistance.

